# A Biomarker-Integrated Day 7 Response Reliably Predicts Outcomes in Minnesota Standard Risk Acute GVHD

**DOI:** 10.64898/2026.10.28.26366649

**Authors:** Ioannis Evangelos Louloudis, Amin Alousi, Tingyu Wang, Marcio Diniz, Daniela Weber, Francis Ayuk, Hannah Choe, Ingrid Vasova, Yi-Bin Chen, Muna Qayed, Ryotaro Nakamura, Hassan Alkhateeb, Aaron M Etra, Matthias Wölfl, Ran Reshef, Elizabeth Hexner, Carrie L Kitko, Paibel Aguayo-Hiraldo, Muhammad Ali, Peter Bader, Chantiya Chanswangphuwana, Matthias Eder, Sabrina Kraus, Margaret L MacMillan, John Magenau, Fotios V Michelis, Timothy Olson, Joseph Pidala, Pavan Reddy, Matthias Stelljes, Robert Zeiser, Yu Akahoshi, Janna Baez, Gilbert Eng, Sigrun Gleich, Nikolaos Katsivelos, George Morales, Tara Sebastian, Nikolaos Spyrou, Rachel Young, William J Hogan, James L M Ferrara, John E Levine

**Author notes:** These authors contributed equally.

## Abstract

Acute graft-versus-host disease (GVHD) response is conventionally assessed 28 days after treatment initiation, although treatment escalation often occurs earlier. In Minnesota standard risk acute GVHD, day 7 clinical response alone provides limited prognostic discrimination because many patients without early improvement subsequently respond without additional therapy. We hypothesized that incorporating serum GVHD biomarkers into a day 7 response assessment would improve early prognostic stratification. We studied 790 patients with Minnesota standard risk GVHD treated with systemic therapy. A MAGIC observational cohort was randomly divided into training (n=294) and validation (n=308) cohorts, and an independent interventional trial cohort (n=188) served as external validation. A classification and regression tree integrated day 7 clinical response with Ann Arbor biomarker scores to predict 6-month non-relapse mortality (NRM). The integrated model reclassified approximately 20% of patients, identifying clinical non-responders with low-risk biomarkers who had favorable outcomes and clinical responders with high-risk biomarkers who remained at increased risk. In the training cohort, the integrated model improved prediction of 6-month NRM compared with day 7 clinical response alone (AUC, 0.76 vs 0.63; P=.002) and performed comparably to conventional day 28 clinical response (AUC, 0.70). These findings were reproduced in both validation cohorts. Among day 7 clinical non-responders, biomarker status identified biologically favorable slow responders and also stratified outcomes among patients who received second-line therapy. A biomarker-integrated day 7 response assessment improves early prognostic stratification in Minnesota standard risk acute GVHD and may provide an earlier, more objective endpoint for clinical decision-making and future therapeutic trials.

## INTRODUCTION

Acute graft-versus-host disease (GVHD) remains a major cause of morbidity and non-relapse mortality (NRM) following allogeneic hematopoietic cell transplantation (HCT)^1,2^. Although systemic corticosteroids remain standard first-line therapy^3^, outcomes vary substantially according to disease severity at presentation, with response rates of approximately 40% in Minnesota high risk disease and 70% in standard risk disease^4,5^.

In clinical trials, overall response (OR) at day 28 is conventionally used as a surrogate endpoint for long-term NRM and survival^6^. In practice, decisions to initiate second-line treatment due to lack of response are often made before day 28 and rely heavily on subjective clinical judgment^7–10^, underscoring the need for earlier and more objective assessment tools^6,11,12^. A recent large analysis demonstrated that a symptom-only based assessment, the clinical response on day 7, predicted NRM and survival as well as waiting to assess response at day 28 among patients with severe symptoms at treatment initiation (Minnesota high risk GVHD)^13^. In contrast, day 7 response assessment was inferior to day 28 assessment in Minnesota standard risk GVHD. This limitation largely reflects the presence of “slow responders”, i.e., patients without clinical improvement at day 7 who subsequently achieve response by day 28 without additional therapy and who experience favorable outcomes. Because slow responders cannot be prospectively distinguished from true treatment failures using clinical criteria alone, treatment escalation before day 28 remains highly variable in practice.

The serum biomarkers ST2 (suppression of tumorigenicity 2) and REG3α (regenerating islet-derived protein 3-alpha), reflect gastrointestinal (GI) crypt injury, a major determinant of acute GVHD outcomes^14–16^. The combination of these two biomarkers generates Ann Arbor (AA) scores that provide prognostic information at GVHD onset and during treatment^5,17–20^, but they have not yet been formally integrated into early response assessment. We hypothesized that integrating AA biomarker scores with day 7 clinical response would provide a more accurate assessment of GI crypt injury and thereby improve early prognostic discrimination to a level comparable to the conventional day 28 clinical assessment. We tested this hypothesis in two independent cohorts: an observational Mount Sinai Acute GVHD International Consortium cohort (MAGIC) and an external validation cohort of interventional trial participants.

## METHODS

### Study Design

This study included two independent cohorts of patients treated for Minnesota standard risk acute GVHD. An observational trial cohort included patients enrolled in the MAGIC database and biorepository who underwent allogeneic HCT between January 2015 and December 2023 and received systemic corticosteroid treatment (≥0.5 mg/kg/d methylprednisolone equivalent, MPE). Eligible patients had complete GVHD staging data at treatment initiation and at days 7 and 28 post-treatment. Clinical data and biospecimens were collected according to a PRoBE (prospective-specimen collection, retrospective-blinded evaluation) design as previously described^21^. The clinical severity of acute GVHD was staged using published criteria^22^.

The interventional trial cohort included patients enrolled in four multicenter primary GVHD treatment trials between 2016 and 2023. Two of the trials were conducted by the Blood and Marrow Transplant Clinical Trials Network (BMT CTN): BMT CTN 1501 was a phase 2 randomized trial comparing sirolimus monotherapy with high-dose corticosteroids for Minnesota standard risk GVHD^23^. BMT CTN 1705 (NCT04167514) was a phase 3 randomized, placebo controlled, trial comparing alpha 1-antitrypsin plus high-dose corticosteroids versus high-dose corticosteroids as primary treatment for clinically significant acute GVHD that included patients with Minnesota high risk (66%) and Minnesota standard risk GVHD with GI involvement (34%). Two other interventional trials were conducted by MAGIC. The first trial evaluated the selective JAK1 inhibitor itacitinib alone as an alternative to systemic corticosteroids for low risk GVHD defined by clinical and biomarker criteria^24^. The second trial tested the combination of high dose corticosteroids and natalizumab for high risk GVHD defined by biomarker criteria and included patients with Minnesota high risk (52%) and Minnesota standard risk GVHD (48%)^25^. Only Minnesota standard risk patients from these trials were included in the analyses.

We excluded patients without a day 7 serum sample and those treated for grade I GVHD given that systemic treatment is not typically recommended and may be harmful^9,26^ (**Figure S1**). In the training cohort, patients who died before day 7 were excluded because their Ann Arbor scores could not be assigned. In the validation cohort, however, these patients were retained and classified as non-responders to better reflect real-world practice.

This study was approved by the Institutional Review Boards or equivalent Ethical Boards of the participating centers.

### GVHD Clinical Response

Complete response (CR) was defined as complete resolution of GVHD symptoms in all target organs. Partial response (PR) was defined as improvement by at least one stage in one or more organs without worsening in any other organ. Non-response (NR) included stable disease, progression in any organ, initiation of additional therapy, or death. CR and PR were combined into a single overall response (OR) category at day 28 according to standard practice, and at day 7 based on prior validation^13^. “Slow responders” were patients with clinical NR at day 7 who were in CR or PR on day 28 without additional therapy. Unless otherwise specified, “clinical response” refers to OR.

### Biomarker determination

Samples were shipped to a central laboratory where ST2 and REG3α were analyzed by enzyme-linked immunosorbent assay in batches as previously described^17,27^. MAGIC algorithm probabilities (MAP) were calculated as a value between 0.001 and 0.999 according to the formula: log[–log(1 – MAP)] = –11.263 + 1.844(log_10_ST2) + 0.577(log_10_REG3α). Ann Arbor (AA) scores were assigned using previously validated MAP thresholds (AA1 < 0.141; 0.141 ≤ AA2 < 0.291; AA3 ≥ 0.291).^28,29^

### Statistical Analysis

We randomly divided the observational trial cohort into training (n=294) and internal validation (n=308) cohorts; the interventional trial cohort (n=188) served as an external validation cohort. The primary endpoint was six-month NRM. Cumulative incidences of NRM and relapse were estimated using competing risks (treating relapse or second HCT as competing events for NRM, and vice versa)^30^. Groups were compared with the Gray test^31^. Overall survival was estimated using the Kaplan-Meier method and compared using the log-rank test^32^. Predictive performance was assessed using time-dependent area under the receiver operating characteristics curve (AUC) for six-month NRM with competing risk^33^. Differences in time-dependent AUCs were tested using an influence function–based Wald test, as implemented in the “compare” function of the timeROC package in R^33^. P-values were adjusted for multiple comparisons using the false discovery rate (FDR) method^34^.

To develop a model that integrated early clinical response and biomarker status, we built a classification tree using OR and Ann Arbor score at day 7 to predict six-month NRM^35^. Receipt of second-line therapy by day 7 was not included in the recursive partitioning process but was classified as NR in all subsequent analyses. To guard against overfitting the decision tree model, we used 10-fold cross-validation to optimize the model’s complexity parameter (i.e., the amount of improvement required to create a new branch); the model was repeatedly trained using 90% of the data and then validated on the remaining 10%. This approach minimized cross-validated error (an estimate of how the model will perform on new data), thus optimizing predictive accuracy. Each terminal node, the final groups into which patients were sorted, was required to contain at least 30 patients, so that the NRM estimate for each group rested on an adequately sized sample. To derive a binary risk stratification, the resulting terminal nodes were aggregated into two groups by hierarchical clustering using Ward’s minimum variance criterion on node-level NRM rates^36^. All analyses were performed in R, version 4.3.3^37^.

## RESULTS

### Patient Characteristics

A total of 790 patients with Minnesota standard risk, grade II-IV acute GVHD who received systemic treatment and had serum samples available 7 days after treatment initiation were included in this analysis. Patients enrolled in the MAGIC observational cohort were randomly assigned into a training cohort (n = 294) and an internal validation cohort (n = 308). An additional 188 patients who participated in BMT CTN or MAGIC interventional clinical trials of GVHD therapy constituted a second, independent validation cohort.

Baseline characteristics were broadly similar across the three cohorts (**Table 1**). The main difference was a higher proportion of pediatric patients in the observational cohorts (15%) than in the interventional cohort (2%) due to the exclusion of children from most interventional trial protocols. Post-transplant cyclophosphamide (PTCy)-based prophylaxis was also more common in the interventional cohort. Target organ involvement was similar across cohorts and represented the full spectrum of Minnesota standard risk GVHD.

**Table 1.**
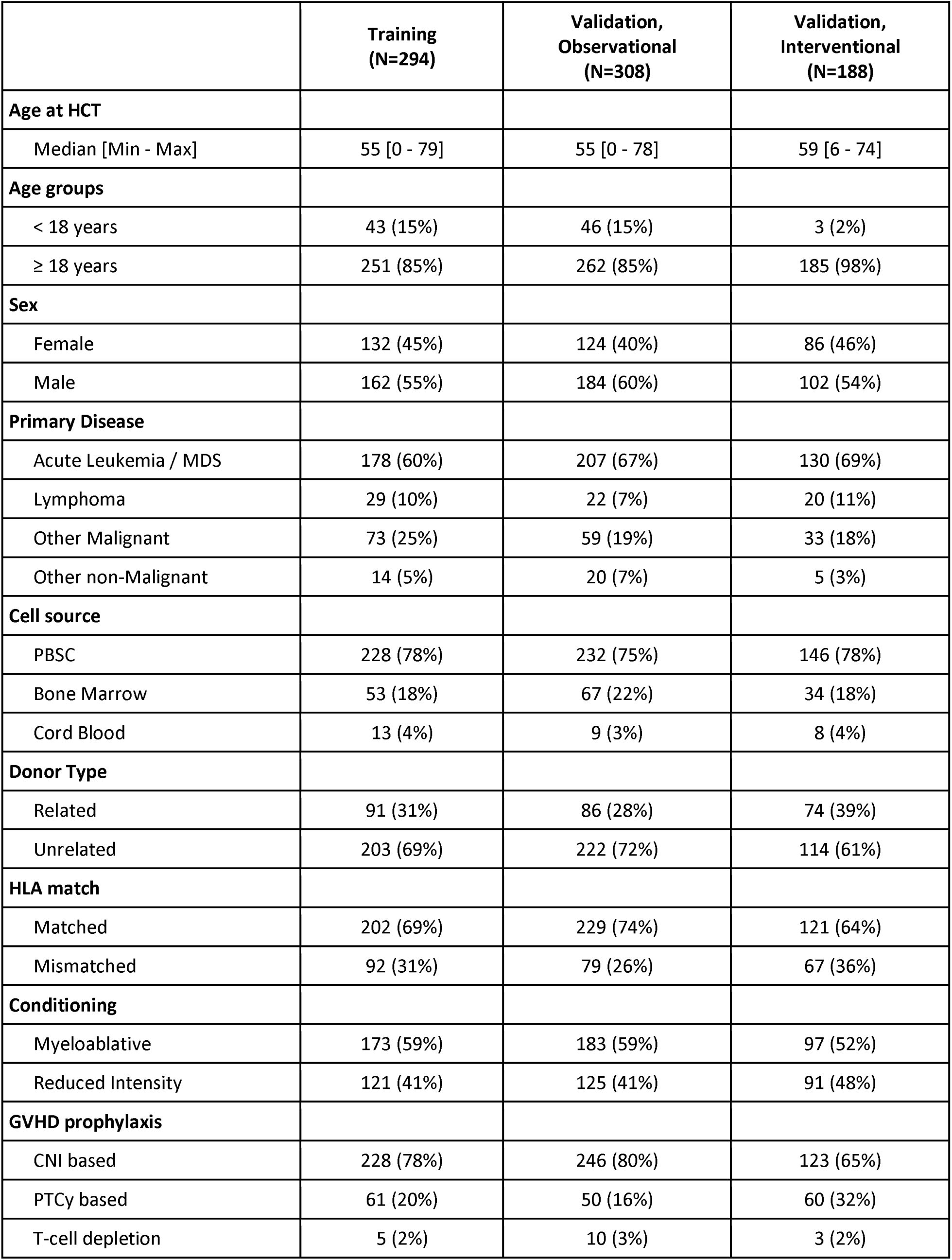
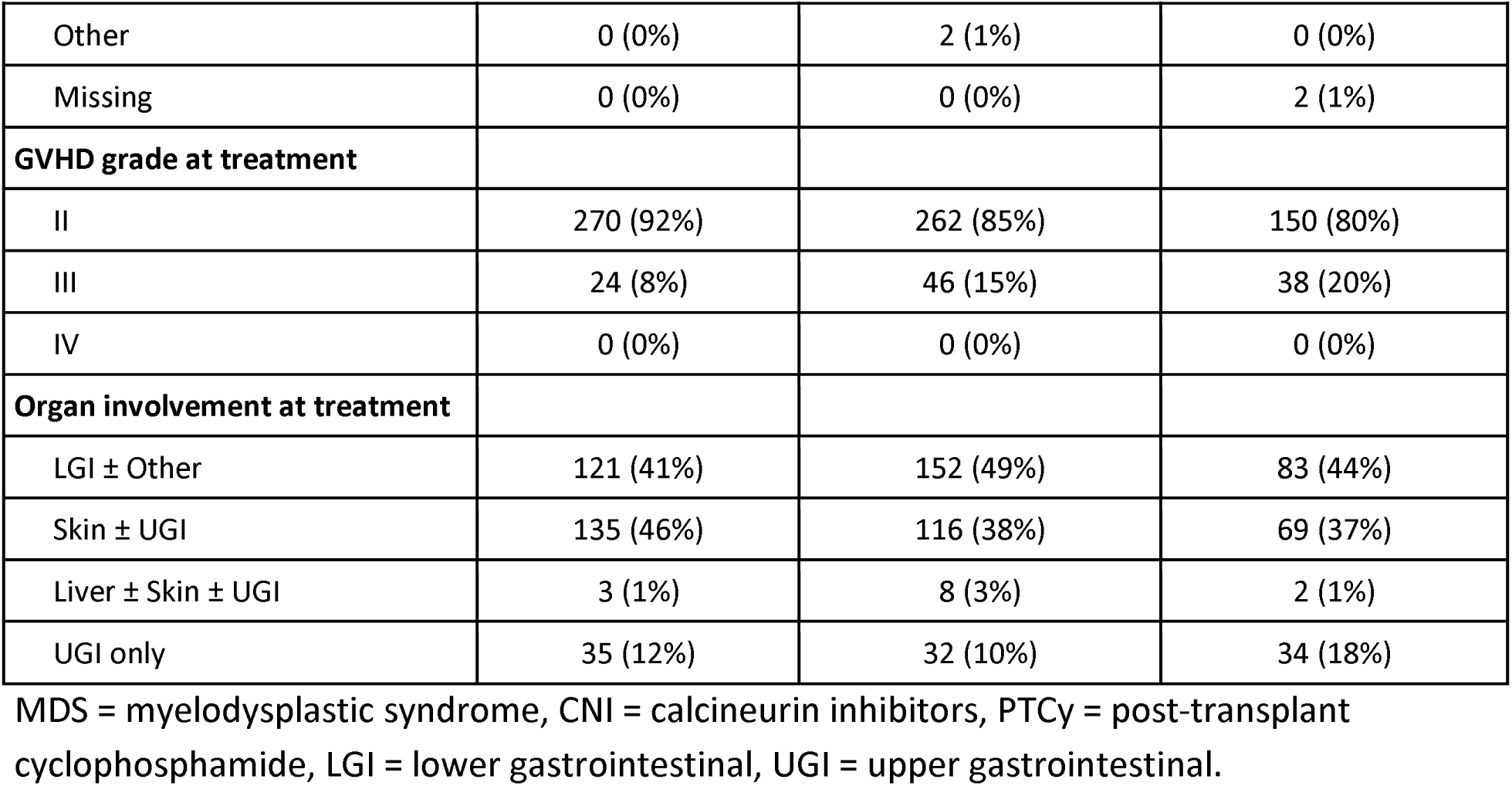
Patient characteristics.

|  | Training<br>(N=294) | Validation,<br>Observational<br>(N=308) | Validation,<br>Interventional<br>(N=188) |
| --- | --- | --- | --- |
| **Age at HCT** |  |  |  |
| Median [Min - Max] | 55 [0 - 79] | 55 [0 - 78] | 59 [6 - 74] |
| **Age groups** |  |  |  |
| < 18 years | 43 (15%) | 46 (15%) | 3 (2%) |
| ≥ 18 years | 251 (85%) | 262 (85%) | 185 (98%) |
| **Sex** |  |  |  |
| Female | 132 (45%) | 124 (40%) | 86 (46%) |
| Male | 162 (55%) | 184 (60%) | 102 (54%) |
| **Primary Disease** |  |  |  |
| Acute Leukemia / MDS | 178 (60%) | 207 (67%) | 130 (69%) |
| Lymphoma | 29 (10%) | 22 (7%) | 20 (11%) |
| Other Malignant | 73 (25%) | 59 (19%) | 33 (18%) |
| Other non-Malignant | 14 (5%) | 20 (7%) | 5 (3%) |
| **Cell source** |  |  |  |
| PBSC | 228 (78%) | 232 (75%) | 146 (78%) |
| Bone Marrow | 53 (18%) | 67 (22%) | 34 (18%) |
| Cord Blood | 13 (4%) | 9 (3%) | 8 (4%) |
| **Donor Type** |  |  |  |
| Related | 91 (31%) | 86 (28%) | 74 (39%) |
| Unrelated | 203 (69%) | 222 (72%) | 114 (61%) |
| **HLA match** |  |  |  |
| Matched | 202 (69%) | 229 (74%) | 121 (64%) |
| Mismatched | 92 (31%) | 79 (26%) | 67 (36%) |
| **Conditioning** |  |  |  |
| Myeloablative | 173 (59%) | 183 (59%) | 97 (52%) |
| Reduced Intensity | 121 (41%) | 125 (41%) | 91 (48%) |
| **GVHD prophylaxis** |  |  |  |
| CNI based | 228 (78%) | 246 (80%) | 123 (65%) |
| PTCy based | 61 (20%) | 50 (16%) | 60 (32%) |
| T-cell depletion | 5 (2%) | 10 (3%) | 3 (2%) |
| Other | 0 (0%) | 2 (1%) | 0 (0%) |
| Missing | 0 (0%) | 0 (0%) | 2 (1%) |
| **GVHD grade at treatment** |  |  |  |
| II | 270 (92%) | 262 (85%) | 150 (80%) |
| III | 24 (8%) | 46 (15%) | 38 (20%) |
| IV | 0 (0%) | 0 (0%) | 0 (0%) |
| **Organ involvement at treatment** |  |  |  |
| LGI ± Other | 121 (41%) | 152 (49%) | 83 (44%) |
| Skin ± UGI | 135 (46%) | 116 (38%) | 69 (37%) |
| Liver ± Skin ± UGI | 3 (1%) | 8 (3%) | 2 (1%) |
| UGI only | 35 (12%) | 32 (10%) | 34 (18%) |

MDS = myelodysplastic syndrome, CNi = calcineurin inhibitors, PTCy = post-transplant cyclophosphamide, LGI = lower gastrointestinal, UGI = upper gastrointestinal.

Clinical trials categorize patients as non-responders at the time second line treatment is initiated regardless of clinical response at the time. In both cohorts, second line treatment was initiated earlier and more often after the approval of ruxolitinib for steroid-refractory acute GVHD in 2019^38^ (**Figure S2**). The median time to second line treatment was between 8-14 days.

### Development of the Day 7 Integrated Response

We performed a classification and regression tree (CART) analysis in the training cohort to develop an integrated day 7 response model using 6 month NRM as the endpoint (**Figure 1A**).

**Figure 1.**
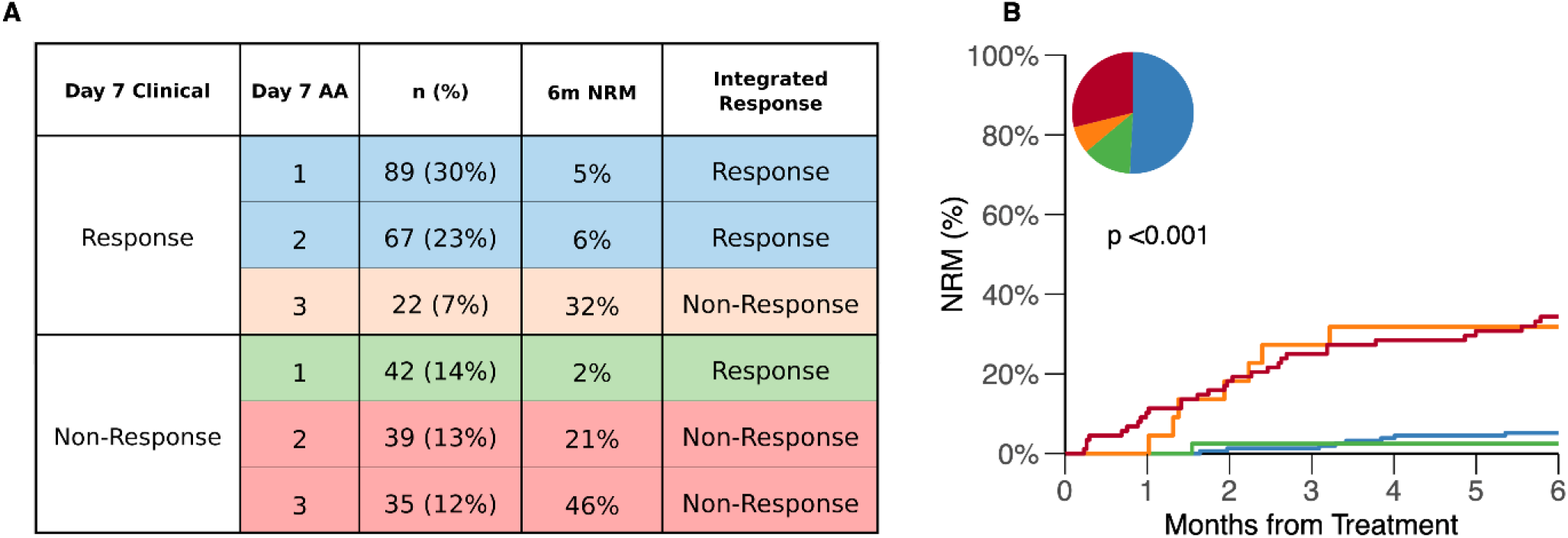
Day 7 integrated model. (A) Classification by day 7 integrated response according to clinical + biomarker criteria. (B) Cumulative incidence of six-month NRM by clinical response and integrated response at day 7. AA = Ann Arbor score; NRM = non-relapse mortality.

We divided each response category into three AA scores to form a total of six strata. Non-responding patients with AA1 scores exhibited very low NRM (2%) and were therefore reclassified as responders. Conversely, responding patients with AA3 scores exhibited high NRM (32%) and were reclassified as non-responders (**Figure 1B**). A total of 21% of patients were reclassified.

We next compared the prognostic performance for NRM of the day 7 integrated model with conventional clinical response assessments at days 7 and 28 in the training cohort. The integrated model yielded greater discrimination between responders and non-responders than day 7 clinical response alone (5% vs 31% compared to 9% vs 21%), and comparable discrimination to the conventional day 28 response (8% vs 33%) (**Figure S3**). Consistent with this finding, the area under the receiver operating characteristics curve (AUC) for predicting six-month NRM was significantly higher for the day 7 integrated model than for day 7 clinical response alone (0.76 vs 0.63; P = .002) and was numerically higher but did not significantly differ from the AUC of the day 28 clinical response model (0.70; P = .176). These findings were reflected in the performance metrics (i.e., sensitivity, specificity, positive and negative predictive value) of the three models **(Table S1**).

### Validation of the Day 7 Integrated Response

We then evaluated model performance in two independent cohorts by comparing three predictors of NRM: day 7 clinical response, day 7 integrated response (clinical plus biomarkers), and the current standard, day 28 clinical response. As expected based on our previous study^13^, day 7 clinical responders had lower NRM than non-responders, but the separation between groups was modest, particularly in the interventional cohort where the difference was not statistically significant (**Figure 2A**). Integration of AA scores reclassified approximately 18% of patients, similar to the training cohort. The integrated model created a highly significant difference between response groups in both cohorts (**Figure 2B**). The proportion and outcomes of the groups by day 7 integrated criteria closely mirrored those of the day 28 clinical response model (**Figure 2C**).

**Figure 2.**
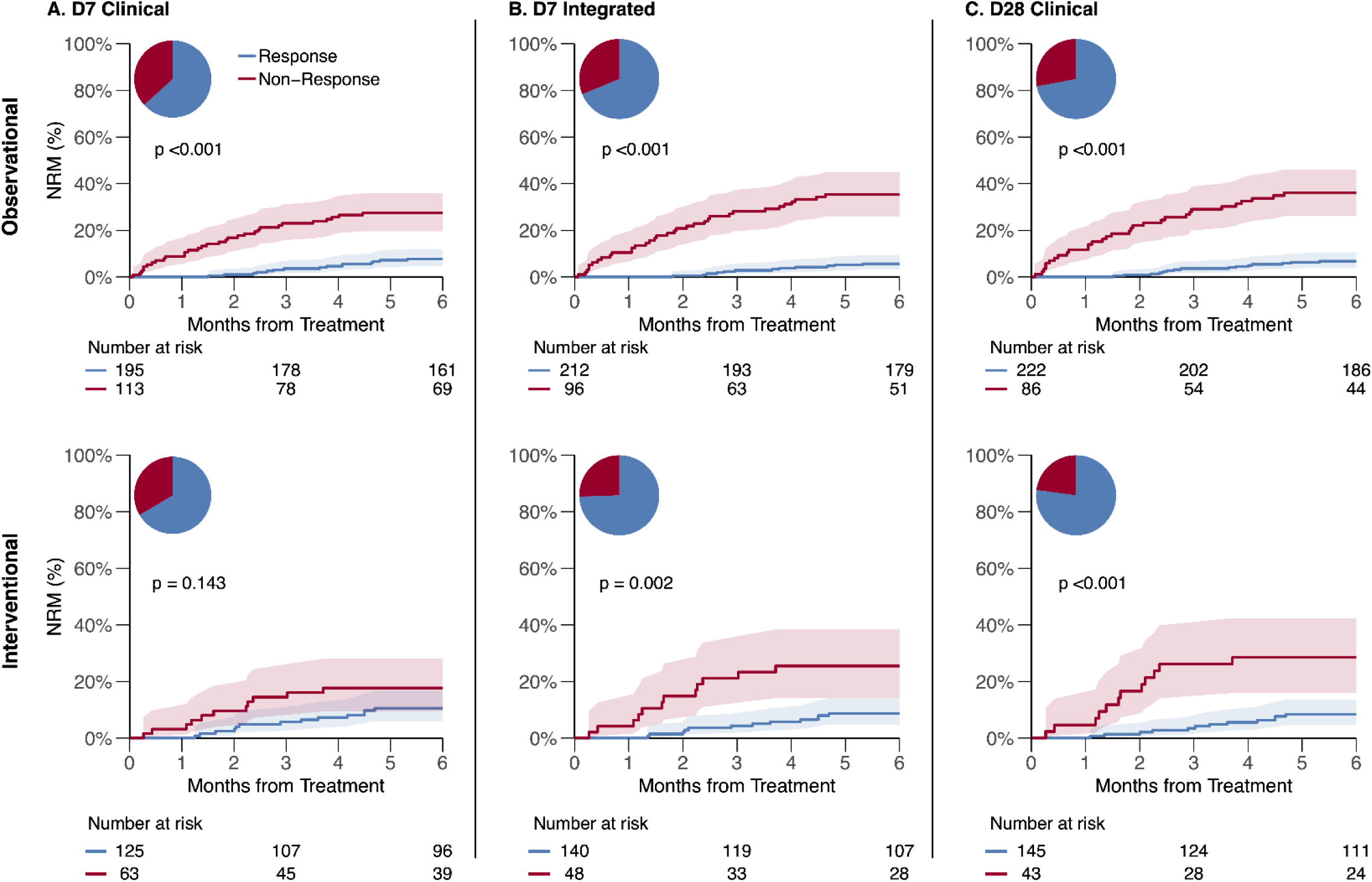
Cumulative incidence of six-month NRM by response models (validation cohorts). (A) D7 Clinical — Observational: 8% vs 27%, p <0.001; Interventional: 11% vs 18%, p = 0.143. (B) D7 Integrated — Observational: 6% vs 35%, p <0.001; Interventional: 9% vs 25%, p = 0.002. (C) D28 Clinical — Observational: 7% vs 36%, p <0.001; Interventional: 8% vs 28%, p <0.001. NRM = non-relapse mortality. Shading corresponds to 95% confidence intervals.

These findings were reflected in the overall predictive performance of the models. In both validation cohorts, the day 7 integrated response achieved higher AUCs for six-month NRM than day 7 clinical response alone and performed comparably to day 28 clinical response. In the observational cohort, AUCs were 0.68, 0.75, and 0.73 for day 7 clinical, day 7 integrated, and day 28 clinical response, respectively; in the interventional cohort corresponding AUCs were 0.57, 0.64, and 0.68. The day 7 integrated response also showed higher sensitivity, specificity, positive predictive value, and negative predictive value than the day 7 clinical response alone in both validation cohorts, with performance metrics similar to those of day 28 clinical response (**Tables 2 and 3**).

**Table 2.** Performance metrics (observational validation cohort)

| Model | ORR | Sensitivity | Specificity | PPV | NPV |
| --- | --- | --- | --- | --- | --- |
| D7 Clinical | 63% | 67% | 69% | 28% | 92% |
| D7 Integrated | 69% | 74% | 77% | 36% | 94% |
| D28 Clinical | 72% | 67% | 79% | 36% | 93% |

ORR = overall response rate, PPV = positive predictive value, NPV = negative predictive value

**Table 3.** Performance metrics (interventional validation cohort)

| Model | ORR | Sensitivity | Specificity | PPV | NPV |
| --- | --- | --- | --- | --- | --- |
| D7 Clinical | 66% | 46% | 69% | 18% | 90% |
| D7 Integrated | 74% | 50% | 79% | 26% | 91% |
| D28 Clinical | 77% | 50% | 82% | 30% | 92% |

ORR = overall response rate, PPV = positive predictive value, NPV = negative predictive value

### Reclassification of Day 7 Clinical Non-Responders by Biomarker Risk

To understand the improved performance of the integrated model, we examined patients classified as clinical non-responders at day 7. By day 7, approximately 25% of patients had already experienced an event defining treatment failure (death or initiation of second-line therapy; **Supplemental Table 2**). The remaining 130 day 7 clinical non-responders from the validation cohorts were pooled for analysis. Clinical features associated with poor prognosis remained important within this subgroup. Patients with lower gastrointestinal involvement had worse NRM than those without lower GI involvement as did patients with progressive GVHD compared to those with stable disease (**Figure 3**). Within each subgroup, however, the AA score further stratified patients according to risk of 6-month NRM (**Table 4**), although the separation among patients with progressive GVHD, while large, did not reach statistical significance due to the small number of patients.

**Figure 3.**
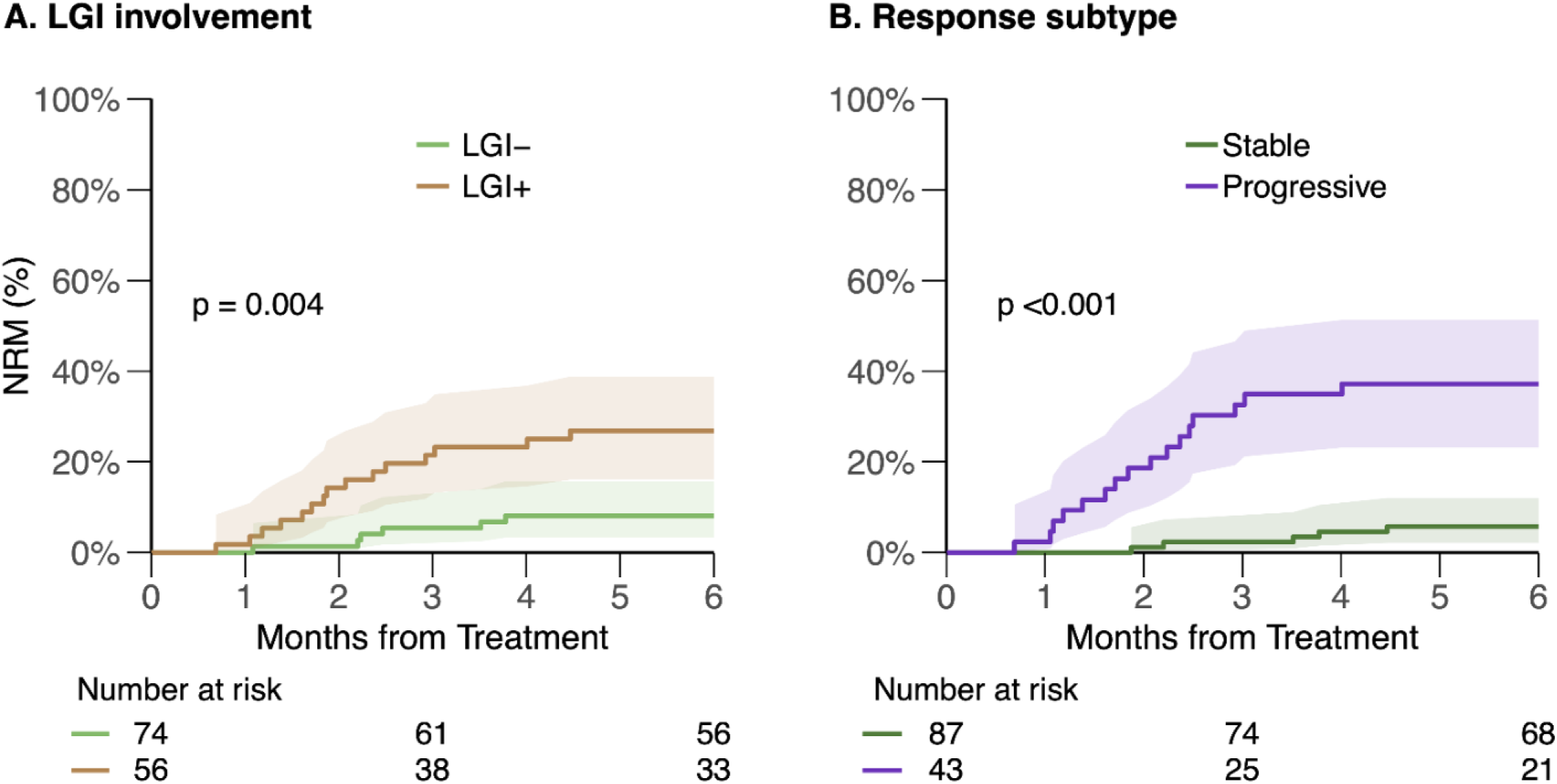
Cumulative incidence of six-month NRM for day 7 clinical non-responders by response subtype and LGI involvement. (A) LGI– vs LGI+: 8% vs 27%, p = 0.004. (B) Stable vs progressive: 6% vs 37%, p < 0.001.

**Table 4.**
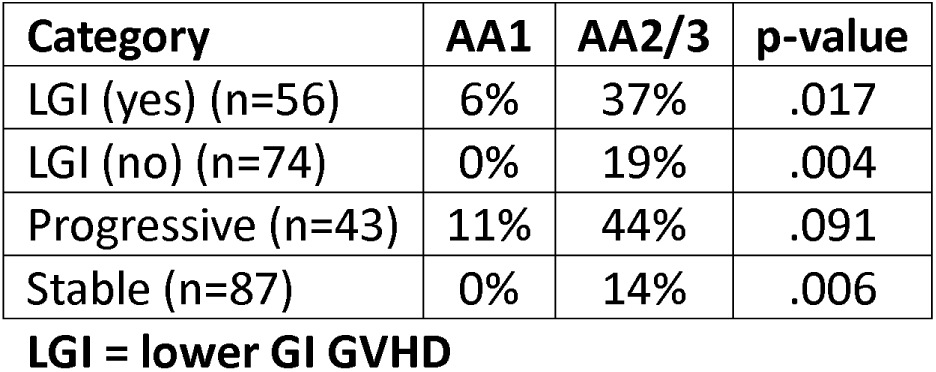
Six-month NRM of day 7 non-responder categories by Ann Arbor scores.

We previously showed that the limited prognostic value of day 7 clinical response in patients with Minnesota standard risk GVHD is largely attributable to patients who fail to respond by day 7 but who subsequently improve without additional therapy (“slow responders”)^13^. Analysis of patients who remained on first-line therapy despite day 7 clinical non-response showed that patients with AA1 biomarkers experienced 0% NRM compared to AA2/3 biomarkers who had 20% NRM (P=.001) (**Figure 4A**). Most patients with AA1 subsequently became slow responders. Serum biomarkers on day 7 similarly stratified patients who initiated second-line therapy before day 28 (AA1 v AA2/3: 8% vs 42% NRM, P=.036) (**Figure 4B**). Notably, outcomes among patients with AA1 biomarkers were similar, regardless of whether second-line therapy was administered, although the number of patients was insufficient to evaluate outcomes according to specific second-line therapies such as ruxolitinib.

**Figure 4.**
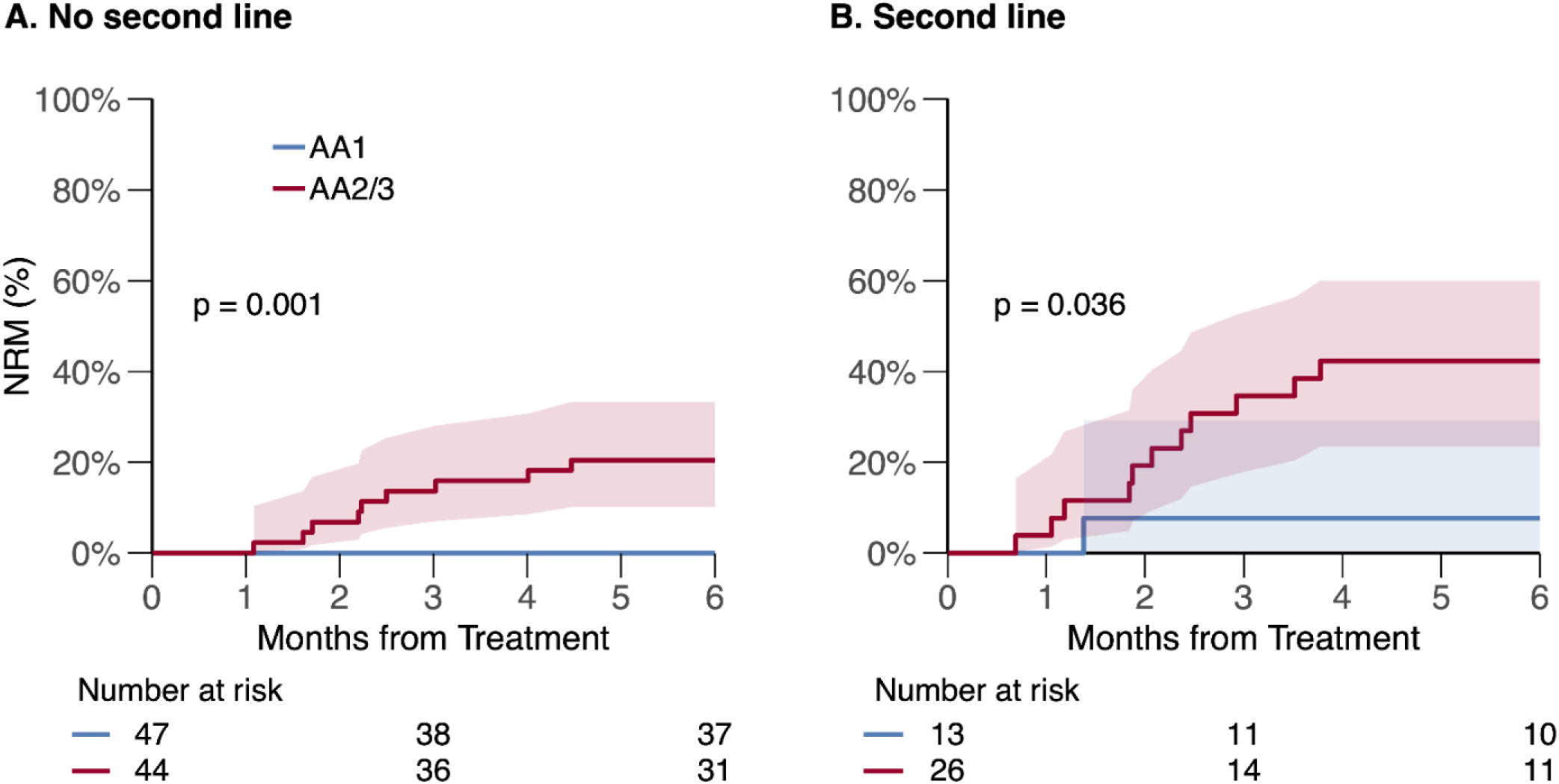
Cumulative incidence of six-month NRM for day 7 clinical non-responders by day 7 Ann Arbor scores. (A) No second line AA1 vs AA2/3: 0% vs 20%, p = 0.001. (B) Second line AA1 vs AA2/3: 8% vs 42%, p = 0.036.

### Overall Survival and Relapse

Six-month relapse rates were similar between response groups in both validation cohorts for all three assessment methods (**Figure 5**). Accordingly, differences in six-month overall survival (OS) closely paralleled those observed for NRM (**Figure 6**). Survival by day 7 integrated response and day 28 clinical response were comparable in both the observational (91% vs 60% and 90% vs 59%, respectively) and interventional cohort (86% vs 70% and 86% vs 69% respectively).

**Figure 5.**
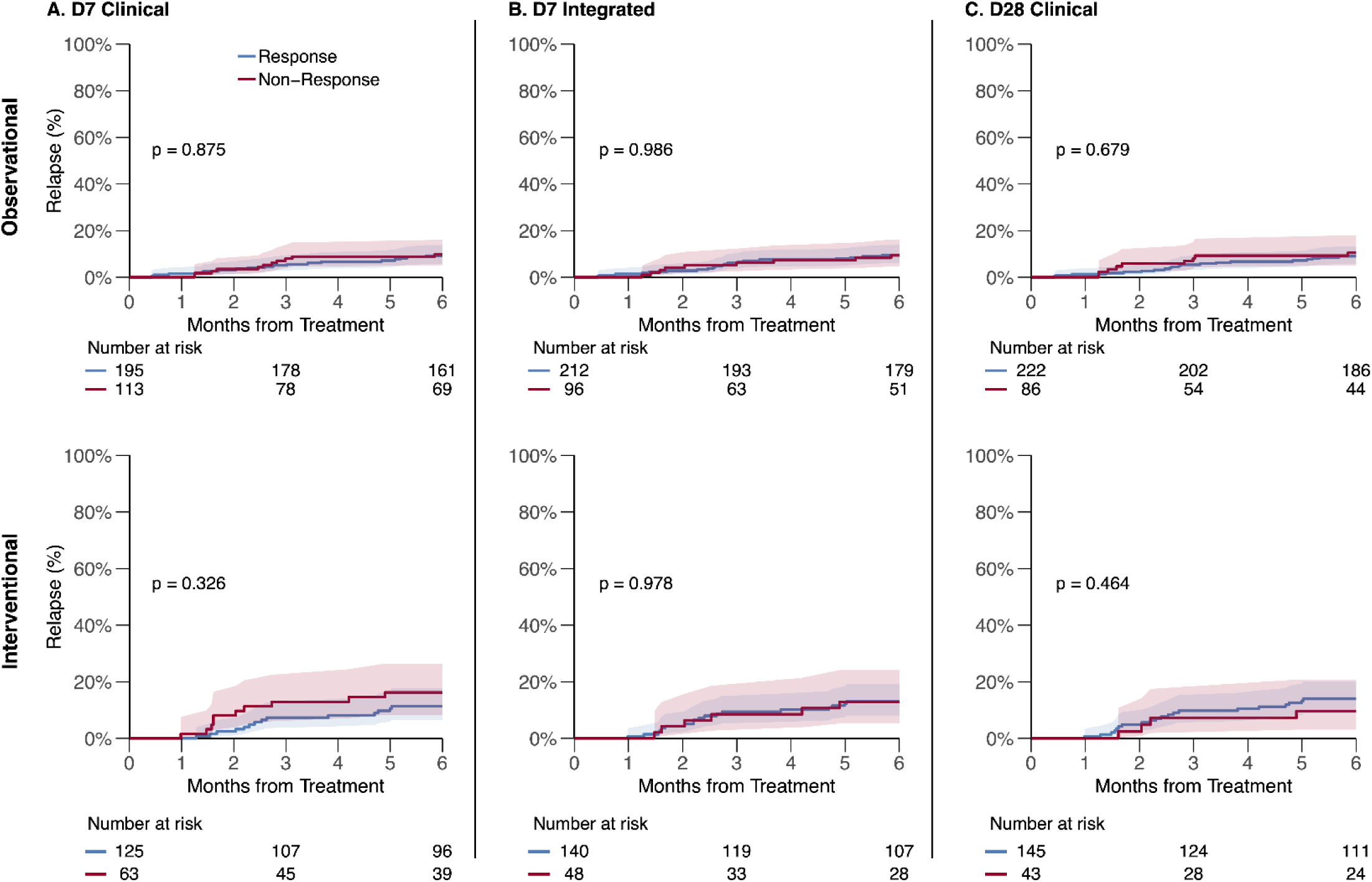
Cumulative incidence of relapse at six months by response models (validation cohorts). (A) D7 Clinical — Observational: 9% vs 10%, p = 0.875; Interventional: 11% vs 16%, p = 0.326. (B) D7 Integrated — Observational: 9% vs 9%, p = 0.986; Interventional: 13% vs 13%, p = 0.978. (C) D28 Clinical — Observational: 9% vs 10%, p = 0.679; Interventional: 14% vs 10%, p = 0.464. Shading corresponds to 95% confidence intervals.

**Figure 6.**
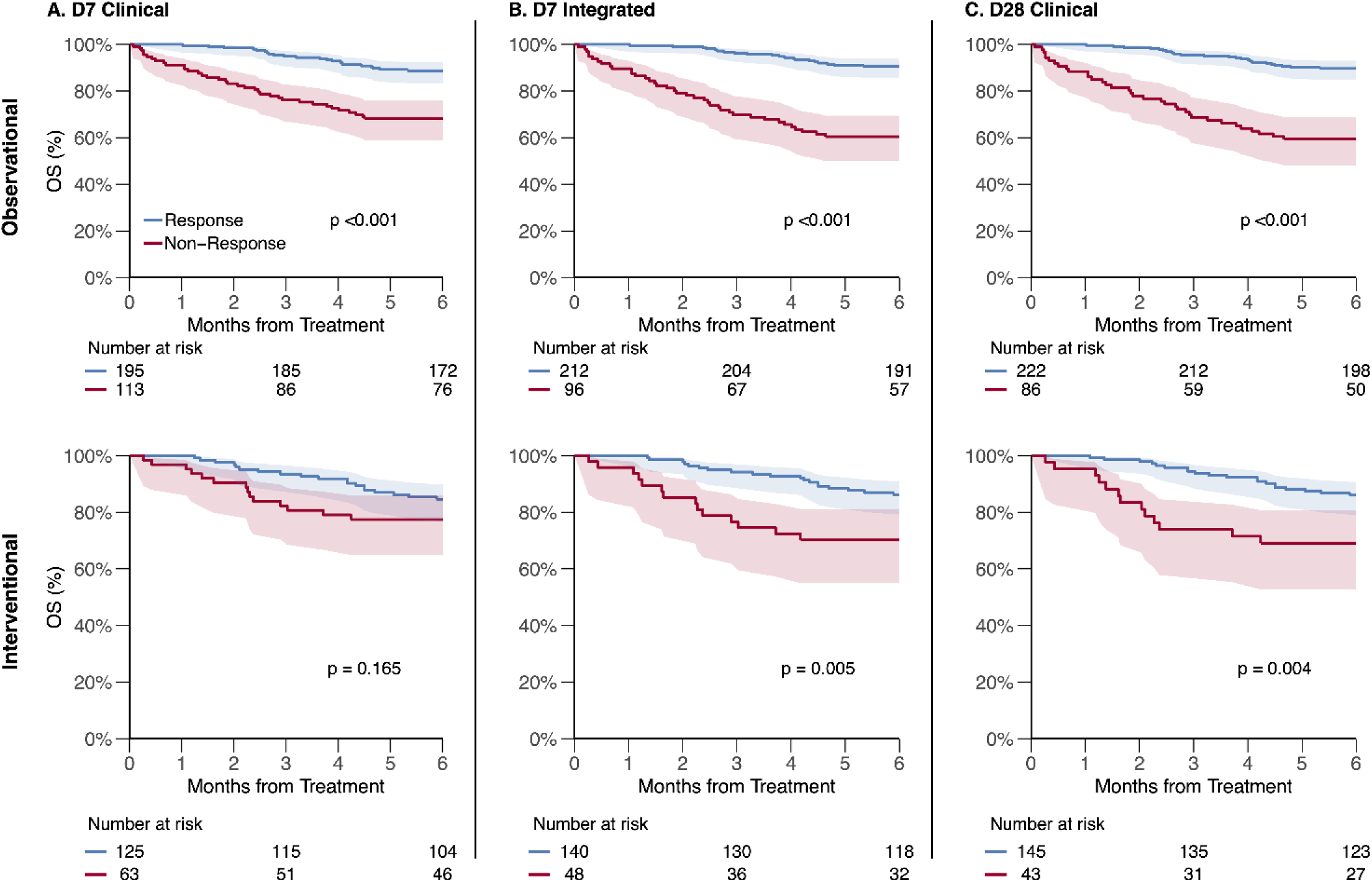
Overall survival at six months by response models (validation cohorts). (A) D7 Clinical — Observational: 89% vs 68%, p <0.001; Interventional: 85% vs 77%, p = 0.165. (B) D7 Integrated — Observational: 91% vs 60%, p <0.001; Interventional: 86% vs 70%, p = 0.005. (C) D28 Clinical — Observational: 90% vs 59%, p <0.001; Interventional: 86% vs 69%, p = 0.004. Shading corresponds to 95% confidence intervals.

## DISCUSSION

This study demonstrates that incorporation of biomarker status into day 7 response assessment improves early prognostic discrimination in Minnesota standard risk acute GVHD. Compared with clinical response alone, the integrated approach more accurately predicted six-month NRM and performed similarly to the conventional day 28 assessment. The findings were reproduced in two independent validation cohorts representing both observational and interventional settings. Approximately one in five patients was reclassified with the addition of biomarkers, notably identifying a substantial subset of clinical non-responders with favorable outcomes and low risk biomarkers as well as a smaller group of apparent responders with high-risk biomarkers. Together, these results indicate that a biomarker-integrated day 7 assessment can provide clinically meaningful risk stratification before most treatment modifications occur. By evaluating response before therapeutic adjustments are made, this approach could reduce the influence of physician behavior on response classification and provide a more objective foundation for treatment decisions.

These findings address a major limitation of early response assessment in Minnesota standard risk acute GVHD. In a recent study of patients with Minnesota high-risk disease, day 7 clinical response was an actionable prognostic marker with predictive performance comparable to day 28 response^13^. In standard risk disease, however, day 7 clinical assessment alone was less informative because approximately half of patients without an early clinical response subsequently improved by day 28 without additional therapy (“slow responders”) and experienced relatively low NRM. In the current analysis, the serum biomarkers stratified these slow responders into biologically distinct groups with divergent outcomes. Such stratification supports incorporation of a biomarker assessment, as it identifies patients likely to experience a delayed recovery.

The biologic rationale is also consistent with current understanding of acute GVHD. The AA score reflects circulating ST2 and REG3α, which are associated with gastrointestinal crypt injury rather than symptom severity alone^17^. Early after treatment initiation, clinical manifestations may not fully reflect the underlying trajectory of tissue injury. Persistent symptoms may coexist with epithelial recovery in some patients, whereas apparent clinical improvement may occur despite ongoing tissue injury in others. The reclassification of some clinical non-responders into a favorable-risk group, and of a smaller proportion of clinical responders into a less favorable-risk group, is consistent with this framework. These findings also align with prior work showing that biomarkers are particularly informative within Minnesota standard risk populations and that GI epithelial injury remains a major determinant of GVHD outcomes^39^.

Several limitations warrant consideration. Because initiation of second-line therapy contributed to the definition of non-response, physician treatment decisions influenced response classification. Additionally, an early integrated response may be less informative for therapies whose effects, particularly on GVHD biomarkers, are not expected within the first week of treatment. Third, the interventional validation cohort was relatively small, limiting the precision of some analyses. Recipients of PTCy-based prophylaxis were underrepresented relative to current transplant practice, which may limit generalizability, although broader adoption of PTCy appears to have shifted GVHD presentation towards less severe phenotypes^40^. Additionally there were too few pediatric patients to support a meaningful subgroup analysis, and patients treated for grade I acute GVHD were not included, although such treatment is not uncommon in routine practice^26^. Finally, initial treatment was heterogeneous. Although a minimum starting steroid dose was required, dosing was variable, and cumulative steroid exposure could not be evaluated as a predictor of long-term outcomes. Some patients also received alternatives to steroid therapy, including sirolimus^23^ and itacitinib^24^, that reflected the clinical trial landscape, but introduced additional heterogeneity.

From a clinical standpoint, these results support day 7 as a potentially actionable assessment timepoint in Minnesota standard risk acute GVHD when biomarker data are available. For patients without a clinical response at day 7, a low biomarker score identifies those likely to respond and for whom continued observation without escalation of therapy is reasonable. The observation that patients with low-risk biomarker profiles had similar outcomes regardless of second-line therapy is limited by sample size but raises the possibility that some patients may be spared unnecessary additional immunosuppression. Importantly, biomarker-based risk stratification also remained predictive in the setting of treatment escalation. Biomarker status discriminated outcomes in patients who initiated second line treatment after day 7, suggesting that the underlying disease biology rather than treatment escalation was a major determinant of outcomes although we were unable to analyze outcomes by specific second line treatments such as ruxolitinib. These findings are consistent with a prior study that demonstrated that ruxolitinib was superior to other second line treatments but less effective for patients with high biomarkers than low biomarkers at its initiation^41^. Together with prior studies in Minnesota high-risk disease these data support a severity-adapted approach to early response assessment across the clinical spectrum of acute GVHD. Whether this strategy can improve outcomes when applied prospectively will need to be assessed in a future clinical trial.

These findings may also be relevant to trial design. Day 28 overall response remains the accepted endpoint in most studies of primary therapy, but it is influenced by investigator decisions regarding additional treatment and may not fully capture biologic disease control. In current practice, treatment modification often occurs before day 28, particularly in the setting of available second-line agents. In this context, the observation that day 7 integrated response yields similar response proportions, survival outcomes, and overall predictive performance to day 28 clinical response in both validation cohorts supports its potential role as an earlier surrogate endpoint that could provide an earlier and more objective measure for risk stratification or response assessment. Such a candidate surrogate endpoint would require prospective validation in interventional studies.

In conclusion, biomarker-integrated response assessment at day 7 improved early prognostic discrimination in Minnesota standard risk acute GVHD and performed comparably to conventional day 28 clinical assessment. The principal advantage of this approach was prospective identification of biologically favorable slow responders among patients without early clinical improvement, while also identifying a smaller subset of apparent responders with persistently high-risk biology. Together with prior findings in Minnesota high-risk disease, these data support a severity-adapted framework for earlier treatment decision-making and future trial design.

## Data Availability

All data produced in the present study are available upon reasonable request to the authors.

## Authorship Statement

I.E.L designed the study, collected the clinical data, conducted the statistical analysis, and wrote the manuscript; M.D. advised statistical methods, reviewed, and revised the manuscript, T.W., N.K., and N.S. collected the clinical data, advised statistical methods, reviewed, and revised the manuscript; A.A., D.W., F.A., H.C, I.V., Y.B.C., M.Q., R.N., H.A., A.M.E., M.W., R.R., E.H., C.L.K., P.A.H., M.A., P.B., C.C., M.E., S.K., M.L.M.. J.M., F.V.M., T.O., J.P., P.R., M.S., R.Z collected the clinical data, reviewed, and revised the manuscript; Y.A., J.B., G.E., S.G., T.S., and R.Y. collected and reviewed the clinical data; G.M. conducted laboratory analyses, reviewed, and revised the manuscript, J.E.L. and J.L.M.F. designed the study, interpreted data, advised methods, reviewed, and revised the manuscript, and organized this project. All authors contributed to the writing of the report and approved the final version of the article.

