## Supplemental Tables and Figures for "A Biomarker-Integrated Day 7 Response Reliably Predicts Outcomes in Minnesota Standard Risk Acute GVHD"

**Supplemental Materials**

**Table S1. Performance metrics (training cohort)**

| Model | ORR | Sensitivity | Specificity | PPV | NPV |
|---|---|---|---|---|---|
| D7 Clinical | 60% | 63% | 64% | 22% | 92% |
| D7 Integrated | 66% | 77% | 74% | 32% | 95% |
| D28 Clinical | 76% | 57% | 82% | 34% | 92% |

ORR = overall response rate, PPV = positive predictive value, NPV = negative predictive value

**Table S2. Categorization of day 7 non-responders in the validation cohort**

| Cohort | Category | N (%) |
|---|---|---|
| All N=176 | Stable | 87 (49%) |
| | Progression | 43 (24%) |
| | 2nd line | 39 (22%) |
| | Death | 7 (4%) |

**Figure S1. Consort diagrams**

**(A) Observational cohort**

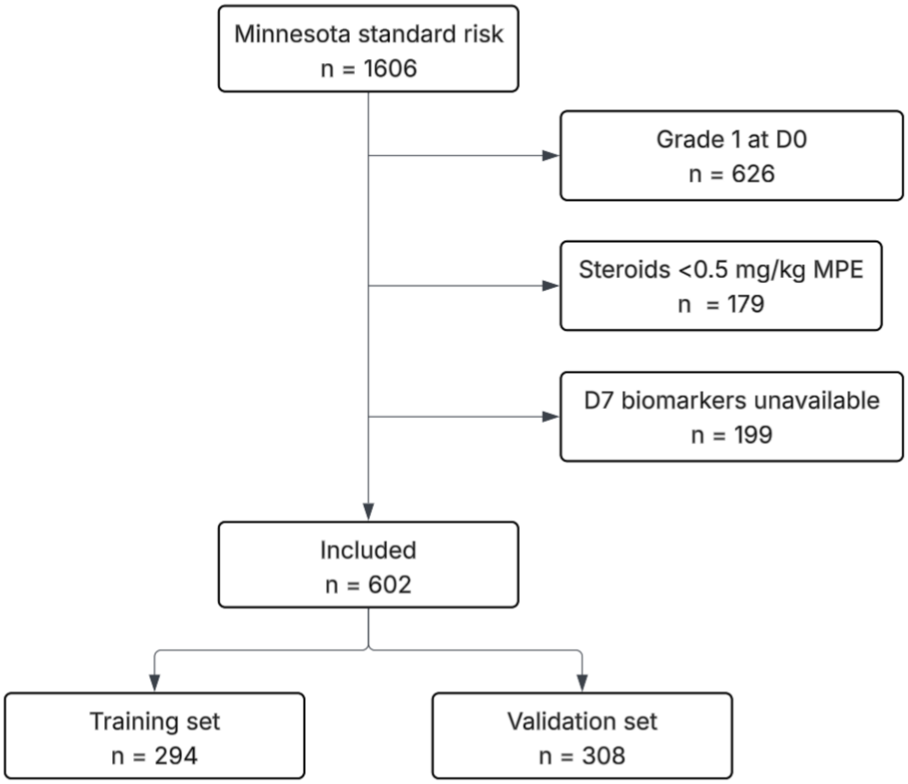

**(B) Interventional cohort**

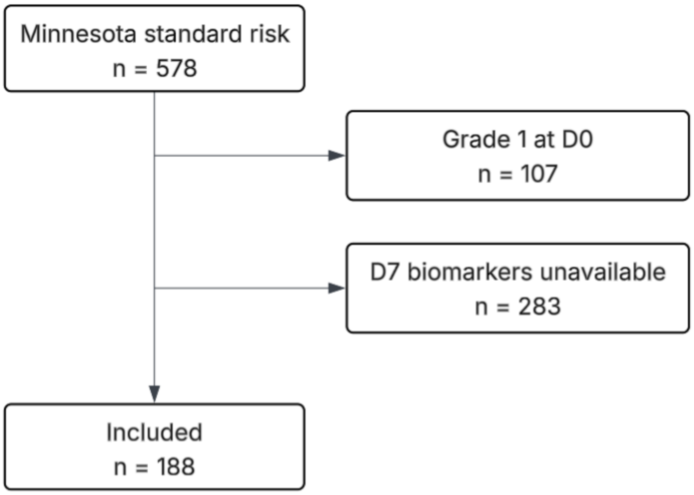

**Figure S2. Second line treatment before day 28 before and after ruxolitinib approval**

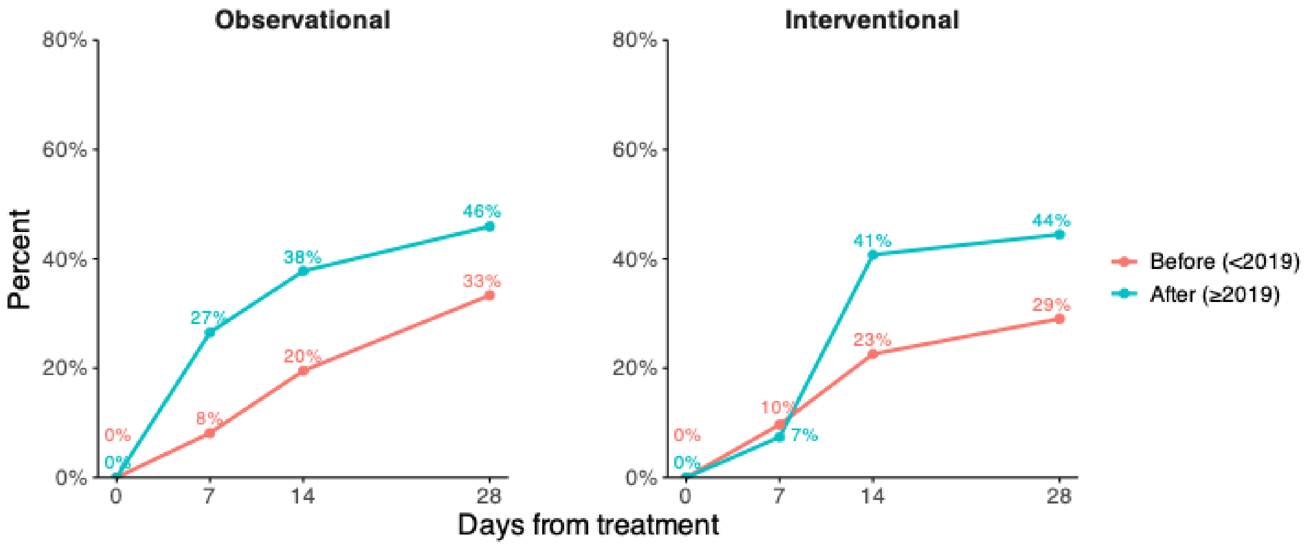

**Figure S3. Cumulative incidence of six-month NRM by response models (training cohort)**

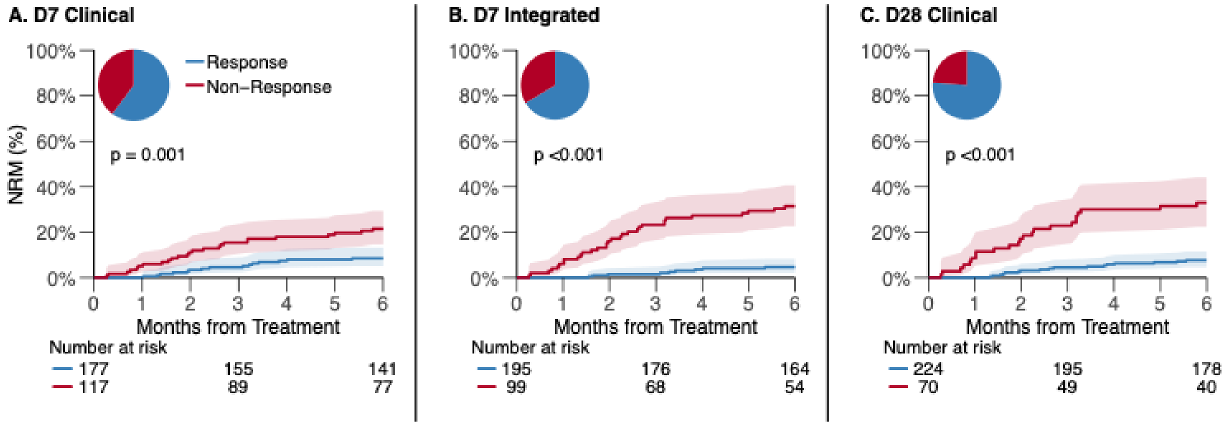

(A) D7 Clinical: 9% vs 21%, p = 0.001. (B) D7 Integrated: 5% vs 31%, p <0.001. (C) D28 Clinical: 8% vs 33%, p <0.001.

NRM = non-relapse mortality. Shading corresponds to 95% confidence intervals.
